# Genetic Architecture and Sample Size Impact Relative Performance of Nonlinear Machine Learning and Standard Polygenic Risk Scores

**DOI:** 10.64898/2026.08.29.26361109

**Authors:** Jingqi Zhu, Alexandra Baousi, Andrew P. Morris, Hui Guo

**Author notes:** Senior authors (these authors jointly supervised the work) (Andrew P. Morris, Hui Guo).

## Abstract

Standard polygenic risk scores (PRSs) are constructed based on additive genome-wide association study summary statistics. Nonlinear machine learning methods have been increasingly applied to construct PRSs from individual-level data, aiming to improve predictive performance over standard PRSs through modelling nonadditive genetic effects. However, their superiority across studies has been inconsistent. The conditions under which they provide meaningful improvements remain unclear. We combined theory, simulations and a real-world application to investigate when two widely used nonlinear machine learning methods, random forest and XGBoost, outperform standard PRSs. Theoretically, we showed that standard PRSs can implicitly capture some genetic variance attributable to nonadditive genetic effects through their contributions to marginal SNP effects. Although nonlinear models have a higher theoretical potential, their bias-variance trade-off can limit predictive gains at finite sample sizes. Simulations showed that XGBoost outperformed standard PRSs only when the genetic architecture involves a large proportion of interaction genetic variance concentrated across relatively few interactions and training sample sizes are large. Random forest consistently underperformed standard PRSs. In risk prediction of ischemic heart disease using UK Biobank data, XGBoost showed little improvement in predictive performance over standard PRSs, whereas random forest again performed worse. Together, these findings suggest that nonlinear machine learning do not uniformly outperform standard PRSs; rather, their relative performance depends jointly on genetic architecture and training sample size. Our study helps to reconcile the inconsistent results reported across previous studies and provides a framework for identifying settings in which more complex PRS models are likely to be beneficial.

## 1. Introduction

Genetic variation contributes to individual differences in many complex traits and disease risks, motivating efforts to quantify genetic predisposition for prediction and risk stratification^1^. Current genome-wide association studies (GWASs) has established that most complex traits and diseases are polygenic^2^. Polygenic risk scores (PRSs), which aggregate the effects of single nucleotide polymorphisms (SNPs) across the genome, have become the predominant approach for quantifying an individual’s genetic predisposition to a phenotype.

The PRS is typically constructed as a weighted sum of effect allele counts, where the weights are the marginal SNP effect estimates obtained from an additive GWAS using effect allele counts as the genotype coding. As marginal effect estimates are subject to linkage disequilibrium (LD) and estimation error, modern PRS methods improve predictive performance by adjusting SNP weights using various strategies^3^. For example, clumping and thresholding (C+T)^4–8^ sets the weights of SNPs that fail certain p-value or LD criteria to zero while retaining the weights of the other SNPs unchanged; Lassosum^9^, LDpred^10,11^, SBayesR^12^ and PRS-CS^13^ shrink SNP weights using penalised regression or Bayesian models that account for LD. Despite their different statistical formulations, these methods all build upon additive GWAS summary statistics. We refer to these collectively as standard PRS methods.

In recent years, machine learning methods have been increasingly applied to construct PRSs directly from individual-level genotype data^14^. Linear machine learning methods, often based on penalised regression, such as lasso^15^, ridge regression^16^ and elastic net^17^, retain a linear statistical formulation but jointly model SNPs using individual-level data, without requiring GWAS summary statistics or an external LD reference panel. Nonlinear machine learning methods, such as random forest^18^ and XGBoost^19^, additionally allow complex nonlinear relationships among SNPs to be modelled and have attracted considerable interest. Their application was mainly motivated by the hypothesis that jointly modelling multiple SNPs using nonlinear methods can learn non-additive (dominance and interaction) genetic effects that are not explicitly modelled by standard PRS methods. As current PRSs often explain only a modest proportion of phenotypic variance, substantially lower than heritability estimates^20^, explicitly modelling non-additive genetic effects has been proposed as one promising avenue for improving PRS performance.

However, evidence for the superiority of nonlinear machine learning PRSs over standard PRSs remains inconsistent across existing studies on various phenotypes, with some demonstrating improved predictive performance^14,21–25^ and others reporting little or no advantage^23,25–28^. Importantly, little attention has been paid to the conditions under which nonlinear machine learning methods outperform standard PRS methods. Identifying such conditions is important for guiding methodological choice and determining when the additional individual-level data and computational costs of machine learning methods are justified by improvements in predictive performance.

In this study, we aim to address this gap through theory, simulation and real-data analysis. First, we show that standard PRSs can capture part of genetic variance attributable to interaction effects, and contrast their theoretical predictive limits with those of nonlinear machine learning methods, which have greater flexibility but face additional challenges in learning from finite samples. Second, we perform comprehensive simulations to compare two widely used machine learning methods – random forest and XGBoost – with standard PRSs across different non-additive polygenic genetic architectures and sample sizes on both quantitative and binary phenotypes. A linear machine learning model, elastic net, is included as a benchmark to distinguish the benefit of nonlinear modelling from that of using individual-level data. Finally, we evaluate these methods in the UK Biobank data using ischemic heart disease as the target phenotype to assess whether the theoretical and simulation findings are supported in a real-world setting.

## 2. Methods

### 2.1 Theory

In quantitative genetics, a phenotype *Y* is often modelled as a function of genetic liability *G* and independent environmental noise *E*. For quantitative phenotypes, we consider *Y* = *G* + *E*. For binary phenotypes, we consider a liability-threshold model, *Y* = *I*(*G* + *E* > *t*), where *I* is the indicator function and *t* is the liability threshold.

For simplicity, we first consider a quantitative phenotype and two independent biallelic SNPs with additive and pairwise additive-by-additive interaction genetic effects on *G*

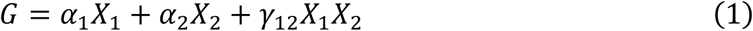

where *X*_1_ and *X*_2_ denote effect allele counts of SNP 1 and 2, respectively. Under Hardy-Weinburg equilibrium, the population marginal effects of SNP 1 on *Y* from an additive GWAS is *β*_1_ = *α*_1_ + 2*p*_2_*γ*_12_, where *p*_2_ is the effect allele frequency of SNP 2. It captures not just the additive genetic effect *α*_1_, but also a proportion of the interaction genetic effect *γ*_12_, where the proportion is determined by the allele frequency of the interaction partner SNP. This occurs because the additive GWAS essentially performs an orthogonal projection of *G* (or equivalently *Y*) onto the linear subspace spanned by the intercept and effect allele count span{1, *X*_1_}. Consequently, the interaction term *γ*_12_*X*_1_*X*_2_ that is correlated with the additive term *α*_1_*X*_1_ contributes to the population marginal effect. Symmetrically, by the orthogonal projection of *G* onto span{1, *X*_2_}, the population marginal effect of SNP 2 on *Y* is *β*_2_ = *α*_2_ + 2*p*_1_*γ*_12_, where *p*_1_ is the effect allele frequency of SNP 1. The genetic liability *G* in Equation 1 can then be decomposed into four mutually orthogonal components

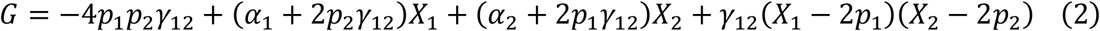

The first three terms on the right-hand side represent the projection of *G* onto the linear subspace spanned by the intercept and all effect allele counts span{1, *X*_1_, *X*_2_} (equivalent to span{1, *X*_1_} + span{1, *X*_2_}) that is estimable by an additive GWAS (Figure 1). The last term is orthogonal to the intercept, *X*_1_ and *X*_2_, and therefore cannot be recovered by any linear predictor in span{1, *X*_1_, *X*_2_}, including standard PRSs constructed from summary-level data and linear machine learning models trained on individual-level data (e.g., elastic net).

**Figure 1.**
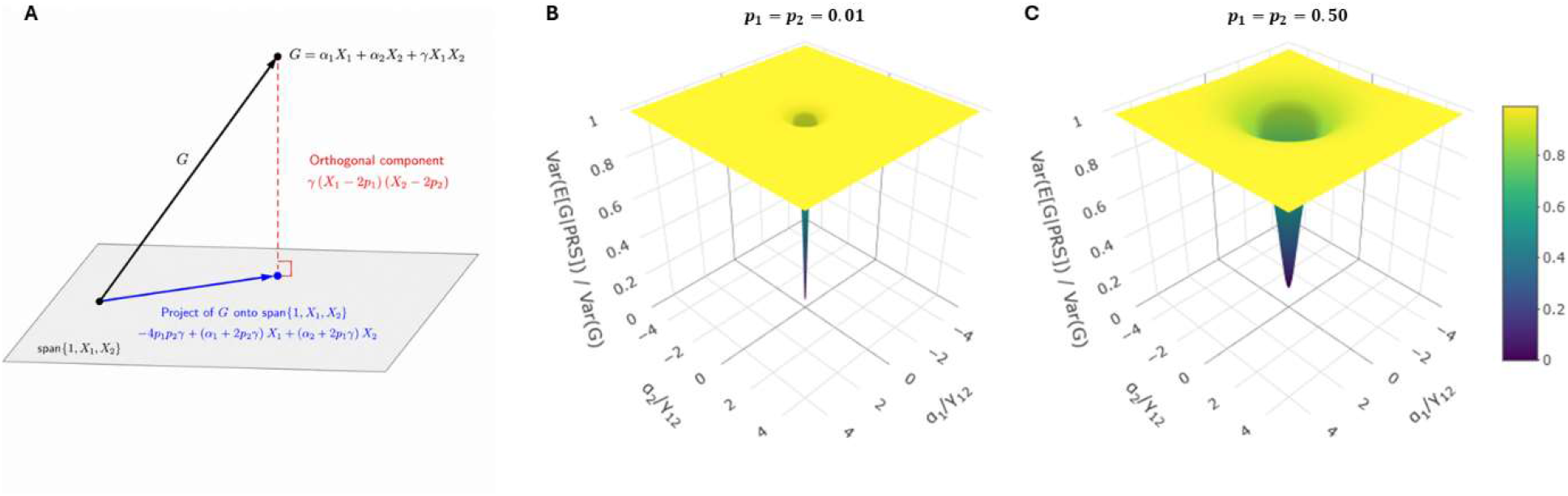
(A) The genetic liability *G* can be decomposed into two components: its projection onto the linear subspace spanned by the intercept and all effect allele counts that is estimable by additive GWASs, and an orthogonal component unrecoverable by linear predictors. (B-C) Theoretical proportion of genetic variance explained by the optimal standard PRS under the two-locus additive-by-additive interaction model, as a function of *α*_1_/*γ*_12_ and *α*_2_/*γ*_12_ with *p*_1_ = *p*_2_ = 0.01, 0.5, respectively. This proportion is minimised to zero at (*α*_1_/*γ*_12_, *α*_2_/*γ*_12_) = (−2*p*_2_, −2*p*_1_), and tends to one as max{*α*_1_/*γ*_12_, *α*_2_/*γ*_12_} → ∞.

Consequently, the optimal standard PRS, defined as the orthogonal projection of the *G* onto span{1, *X*_1_, *X*_2_}, maximises the explained genetic variance over all standard PRSs and can be written as PRS = *β*_1_*X*_1_ + *β*_2_*X*_2_. Note that it is a population-level quantity, rather than an estimated PRS from finite samples, and has explained genetic variance

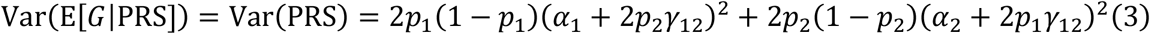

This explained genetic variance can exceed genetic variance due to underlying additive genetic effects 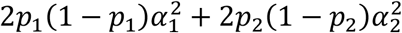, and is exactly the additive genetic variance *V*_A_ in the classical orthogonal decomposition of genetic variance^29^. The unexplained genetic variance

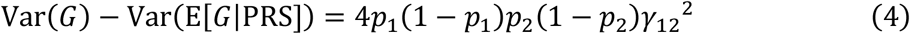

is the interaction genetic variance *V*_I_^30^. The proportion of genetic variance explained by the optimal standard PRS

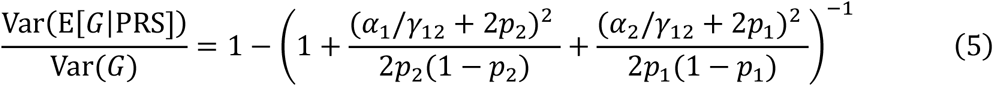

depends on the relationship between the ratios of the additive to additive-by-additive interaction genetic effects ratios *α*_1_/*γ*_12_ and *α*_2_/*γ*_12_, and the effect allele frequencies *p*_1_ and *p*_2_ of the interacting SNPs. It approaches its supremum of one as the magnitude of either |*α*_1_/*γ*_12_| or |*α*_2_/*γ*_12_| tends to infinity; and it attains its minimum when *α*_1_/*γ*_12_ = −2*p*_2_ and *α*_2_/*γ*_12_ = −2*p*_1_ – the interaction-induced components of the marginal SNP effects exactly cancel the underlying additive effects, yielding zero marginal effects. The relationship can be depicted by Figure 1, given equal *p*_1_ and *p*_2_. The optimal standard PRS explains a large proportion of genetic variance across a wide range of *α*_1_/*γ*_12_ and *α*_2_/*γ*_12_ values, with substantial reduction of explained variance confined in a relatively small region that gives rise to weak marginal effects. Moreover, this low-explained-variance region becomes more restricted as the interacting SNPs become rarer.

To extend this argument to a *J*-SNP polygenic model with arbitrary higher-order interactions and LD, we similarly define the optimal standard PRS constructed from *K* SNPs (*K* ≤ *J*) as the orthogonal projection of *G* onto span{1, *X*_1_, …, *X*_*K*_}. Since span{1, *X*_1_, …, *X*_*K*_} ⊆ span{1, *X*_1_, …, *X*_*J*_}, the variance of the standard PRS cannot exceed the variance of the orthogonal projection of *G* onto span{1, *X*_1_, …, *X*_*J*_} which is the additive genetic variance *V*_A_, i.e. Var(PRS) ≤ *V*_A_. The proportion of phenotypic variance explained by the optimal standard PRS *R*^2^(PRS) is bounded above by the additive SNP heritability 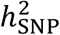. i.e. 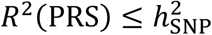.

In practice, as marginal effect estimates from a finite sample GWAS are subject to estimation error and LD, a PRS constructed from these estimates, denoted by 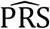, can deviate from the optimal standard PRS, resulting in a lower proportion of explained phenotypic variance, 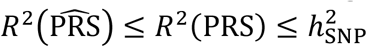. However, some PRSs that explicitly account for LD, such as LDpred, have the desired asymptotic property that, under correctly specified LD and appropriate genetic models, 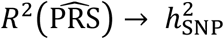 as the GWAS sample size *N* tends to infinity, roughly with a convergence rate *O*(*N*^−1^)^10,20^.

Random forest^18^ and XGBoost^19^ are among the most widely used nonlinear machine learning methods in PRS construction. These tree ensemble methods utilise the nonlinearity of decision trees to capture non-additive genetic effects through decision trees. Specifically, given training data *D*, a tree *f*_tree_ partitions the genotype space into disjoint regions *A*_1_(*D*), …, *A*_*M*_(*D*) via recursive univariate splits and assigns a constant value *c*_*m*_(*D*) to region *A*_*M*_(*D*), and therefore can be written as the sum of a series of indicator basis functions

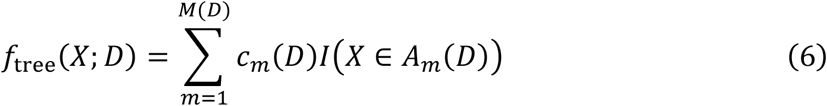

Unlike the standard PRS, trees are capable of directly representing additive and nonadditive genetic effect because they are not subject to any linear subspace. For example, in the aforementioned two-SNP model, the additive genetic effects can be represented with *c*_*m*_*(D*) = 0, *α*_*i*_, 2*α*_*i*_ and *A*_*m*_*(D*) = {*X*_*i*_ = 0}, {*X*_*i*_ = 1}, {*X*_*i*_ = 2}; the additive-by-additive interaction genetic effect can be represented with *c*_*m*_(*D*) = 0, *γ*_12_, 2*γ*_12_, 4*γ*_12_ and *A*_*m*_(*D*) = {*X*_*i*_ = 0 *or X*_*j*_ = 0}, {*X*_*i*_ = *X*_*j*_ = 1}, {(*X*_*i*_ = 1, *X*_*j*_ = 2)*or*(*X*_*i*_ = 2, *X*_*j*_ = 1)}, {*X*_*i*_ = *X*_*j*_ = 2}. In fact, by the universal approximation property, a tree with sufficient depth can approximate the genetic liability arbitrarily well regardless of the underlying LD or the order of genetic interactions^31^. We define such a tree with unrestricted complexity as the population-optimal tree. Compared to the optimal standard PRS, the optimal tree *f*_tree_ = *G* can explain all genetic variance *V*_*G*_, rather than just additive genetic variance *V*_*A*_. The proportion of phenotypic variance explained by the optimal tree *R*^2^(*f*_tree_) can exceed the additive SNP heritability 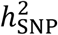 and approach the broad-sense heritability *H*^2^.

However, there is no guarantee on how much the great potential of trees can be realised from finite training data due to the crucial challenge of bias-variance trade-off in machine learning^31^. Increasing tree depth reduces bias in estimating genetic liability by allowing more genetic effects to be represented, but at the cost of greater estimation variance – a small perturbation in the training data can lead to substantially different splits, particularly in terminal regions that contain few observations. Therefore, a tree fitted on finite training data 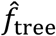 can deviate substantially from the optimal tree *f*_tree_.

Tree ensemble methods were developed precisely to achieve better bias-variance trade-off, thus a more robust approximation of the genetic liability compared to a single tree in PRS construction. For example, random forest averages many trees generated from bootstrapped samples and random SNP subsets^18^, while XGBoost integrates a sequence of shallow trees with explicit regularisation on model complexity^19^. However, it remains no theoretical guarantee on how much the proportion of phenotypic variance explained by random forest and XGBoost from finite training samples, 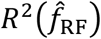 and 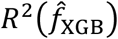, may fall short of their theoretical limit of the broad-sense heritability *H*^2^.

### 2.2 Simulation

We compared the predictive performance of random forest and XGBoost PRSs with standard PRSs, using elastic net as a linear individual-level benchmark, under controlled polygenic architectures comprising both additive and additive-by-additive interaction genetic effects. The description of our simulation study follows the ADEMP framework^32^.

#### Aim

The aim was to determine how the proportion of interaction genetic variance, the number of interaction effects, and sample size affect the relative predictive performance of random forest, XGBoost, elastic net and standard PRSs, with elastic net included as a linear individual-level reference. All models were constructed from the same set of SNPs.

The three factors were chosen to reflect the theoretical considerations outlined above. First, the proportion of interaction genetic variance *λ*_I_ = *V*_I_/*V*_G_ determines the potential predictive gain available to nonlinear models beyond the additive genetic variance – the theoretical limit of standard PRSs. We therefore hypothesised that the relative advantage of random forest and XGBoost over standard PRSs would increase as the proportion of interaction genetic variance *λ*_I_ grew. Second, a given amount of interaction genetic variance may arise from different interaction architectures, ranging from a small number of strong interactions (concentrated interaction architecture) to many weaker interactions (diffuse interaction architecture), which may impact the performance of PRS methods. Third, sample size affects both the accuracy of marginal effect estimates for standard PRSs and the bias–variance trade-off for machine learning PRSs. Finally, by constructing PRSs using the same set of SNPs, we isolated differences in predictive performance attributable to the modelling approach rather than SNP selection.

We additionally considered a linear machine learning method, elastic net, as a benchmark between standard PRSs and nonlinear machine learning methods (random forest and XGBoost). Like the standard PRS, the elastic net PRS is a linear function of the SNP genotypes that belongs to the linear space spanned by all effect allele counts, therefore, the proportion of phenotypic variance it explained is also bounded by the additive SNP heritability. However, unlike the standard PRS, the elastic net PRS is trained directly on individual-level genotype data, as are the random forest and XGBoost PRSs. Therefore, it helps distinguish the contribution of using individual-level data from that of nonlinear modelling.

#### Data-generating mechanisms

We simulated genotype data for *n* individuals and *J* = 200 independent SNPs. The minor allele frequency for each SNP was drawn from a uniform distribution *p*_*j*_ ~ *U*(0.1,0.5), *j* = 1, … *J*. Restricting to common variants avoids rare genotype combinations arising from low-frequency variants, which would incur little interaction genetic variance unless the interaction genetic effects are extremely large. The raw genotype matrix **X** = (*X*)_*n×J*_, *i* = 1, … *n*, was generated from binomial distributions *X*_*ij*_~*Bin*(2, *p*_*j*_) under Hardy-Weinberg equilibrium. The genotype matrix was then standardised 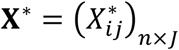 with 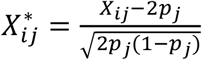 to have expected zero mean and unit variance. The independence between SNPs and the standardisation provide a desired property that all columns of **X**^∗^ and their element-wise products are mutually orthogonal in expectation, thereby allowing the additive and interaction genetic variance to be specified independently. Note that the standardisation was based on population allele frequencies *p*_*j*_ rather than the observed sample mean and variance, which avoided data leakage in the subsequent train-test data split.

The phenotypic model for the quantitative phenotype was **y** = **g** + **e**, where genetic liability **g** was a sum of additive and interaction component vectors **g** = **g**_A_ + **g**_I_ and **e** was a vector for non-genetic residuals. The total phenotypic variance was set to one, and the broad-sense heritability was fixed at *H*^2^ = 0.5, representing a moderately heritable phenotype. This resulted in a total genetic variance of *H*^2^ = 0.5 and residual variance of 1 − *H*^2^ = 0.5. Given the proportion of interaction genetic variance *λ*_I_, the additive and interaction genetic variance were *H*^2^(1 − *λ*_I_) and *H*^2^*λ*_I_, respectively. The additive component was **g**_A_ = **X**^∗^***b***_A_, where **X**^∗^ is the standardised genotype matrix. **b**_A_ was drawn initially from **b**_A_ ~*N*(0, **I**_*J*_) and subsequently rescaled so that **g**_A_ had the target additive genetic variance *H*^2^(1 − *λ*_I_) exactly. For the interaction component **g**_I_, *n*_int_ SNP pairs were sampled without replacement from all 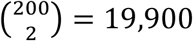 possible pairwise interactions. For the *k*-th sampled pair (*j, j*^’^), 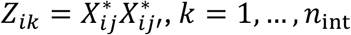. And the standardised interaction genotype matrix was constructed as 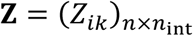. Then, the interaction component was generated as **g**_I_ = **Zd**, where **d** was initially drawn from 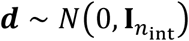 and subsequently rescaled so that **g**_I_ had the target variance *H*^2^*λ*_I_ exactly. The non-genetic noise was generated from **e**~*N*(0, (1 − *H*^2^)**I**_*n*_) and added to genetic liability **g** to create the quantitative phenotype **y**. Finally, the simulated data were randomly split into training (80%) and test (20%) sets.

For binary phenotype, the same procedure was used to simulate the underlying continuous liability. A target prevalence of 0.1 was specified, and the liability threshold was defined as the 90th percentile of the liability distribution in the training set. Individuals with liability exceeding this threshold were assigned as cases, and as controls otherwise. The threshold estimated from the training set was then applied to the test set, ensuring that no information from the test data was used to define the binary outcome.

We varied: *λ*_I_ across {0, 0.1, 0.2, 0.3, 0.4, 0.5}, ranging from entirely additive genetic variance to equal additive and interaction genetic variance components; the number of interacting pairs *n*_int_ across {2, 20, 199}, corresponding to {0.01%, 0.1%, 1%} of all possible pairwise interactions; and the total number of individuals *n* across {10,000, 100,000}, spanning the scale of moderate-size genetic datasets to large-scale biobanks. These give 36 simulation configurations. Each configuration was replicated 100 times to enable Monte Carlo estimation of predictive performance.

#### Targets (Estimands)

The target of the simulation study was the test-set predictive performance of PRSs constructed using different modelling approaches from the same set of SNPs. Of particular interest was the relative predictive performance between the two machine learning approaches and the standard PRS across the simulated genetic architectures and sample sizes.

#### Methods

Since each SNP was predictive of the phenotype by design, all PRS methods were constructed using the full set of 200 SNPs, ensuring that each method had access to the same complete genetic information. The standard PRS was built by first estimating the marginal effect of each SNP via separate univariable linear or logistic regressions using additive genotype coding, similar to an additive GWAS, and then summing the effect allele counts across SNPs weighted by their estimated marginal effects.

Random forest and XGBoost algorithms were applied to the training data using the 200 SNPs as predictors and the phenotype as the outcome. The resulting PRSs were defined as the predicted values for the quantitative phenotype and the predicted probabilities of a case for the binary phenotype. Hyperparameter tuning was performed on training data via 3-fold cross-validation for XGBoost, and via out-of-bag validation for random forest. The best set of hyperparameters for random forest and XGBoost were found using sequential model-based optimisation.

Elastic net jointly estimates all SNP effects within a linear model by combining L1 and L2 penalties. The elastic net PRS was defined as the linear predictor for the quantitative phenotype and the log-odds for the binary phenotype. Hyperparameter tuning was performed on training data via 3-fold cross-validation. Implementation details for hyperparameter tuning of random forest, XGBoost and elastic net are listed in Supplementary Material Table S1.

We further constructed an “oracle reference” with complete knowledge of the true genetic liability. For the quantitative phenotype, the “oracle reference” was defined as the true genetic liability *g*. For the binary phenotype, it was defined as the true conditional probability of disease given the genetic liability *P*(*Y* = 1 | *g*). The “oracle reference” represents the Bayes-optimal genetic predictor and provides the theoretical upper bound on the predictive performance achievable by any PRS method under the simulated genetic architecture.

Standard and elastic net PRSs were run using a single CPU thread, whereas random forest and XGBoost PRSs used 36 CPU cores. All computations were performed on Intel® Xeon® Gold 6230 “Cascade Lake” processors (2.10 GHz) with 192 GB RAM.

#### Performance

Predictive performance was evaluated on the independent test set. For the quantitative phenotype, performance was assessed using the proportion of phenotypic variance explained *R*^2^. For the binary phenotype, performance was evaluated in terms of Nagelkerke *R*^2^ for overall predictive ability, area under receiver operating characteristic curve and precision recall curve (AUROC and AUPRC) for discrimination, and Brier score for calibration. For each simulation scenario, the mean of each performance measure across the 100 simulation replicates was reported together with its Monte Carlo 95% confidence interval.

### 2.3 UK Biobank Data Analysis

We evaluated the same set of PRS methods for predicting ischemic heart disease using the UK Biobank data (application number: 532314). UK Biobank is a large biomedical dataset including genotype and deep phenotype data for approximately 500,000 participants aged 40-69 years recruited between 2006 and 2010^33^. This analysis was performed on the UK Biobank Research Analysis Platform.

Individuals’ ICD-10 codes from the main diagnosis, secondary diagnosis, external diagnosis and cause of death records were mapped to Phecodes (v1.2b1)^34^. Cases were defined as individuals with Phecode 411 (ischemic heart disease), and controls as individuals without Phecode 411 or any of its exclusion Phecodes.

The analysis was restricted to unrelated White British participants to minimise population stratification and prevent information leakage arising from genetic relatedness between the training and test sets. Individuals with sex mismatch or sex chromosome aneuploidy were excluded. The remaining participants were randomly split into training (80%) and test (20%) sets, with stratification by ischemic heart disease status to preserve the case-control ratio in both sets.

An additive GWAS was performed using the training set individuals only. Variant quality control was applied to the Genomics England (GEL)-imputed genotypes using PLINK2^35^ (v2.0.0-a.6.9), retaining SNPs with minor allele frequency (MAF) ≥ 0.005 and MaCH *r*^2^ ≥ 0.8. Association testing was performed using the two-step whole-genome regression method implemented in REGENIE (v4.1)^36^, adjusting for age, sex and the first 10 genetic principal components to minimise population stratification. For Step 1 of REGENIE, the whole genome regression was additionally restricted to array-genotyped SNPs, with the common inversion and major histocompatibility complex (MHC) regions excluded. For Step 2, approximate Firth correction was used as fall-back for variants with p-value less than 0.001 to correct for potential bias due to case–control imbalance and low minor allele counts.

Marginal SNP effect estimates from the additive GWAS were used to construct standard PRSs using C+T. SNPs were LD-clumped using PLINK2 at *r*^2^ < 0.01 within a 1 Mb window and retained using either a genome-wide significance threshold *p* < 5 × 10^−8^ or a more lenient threshold *p* < 1 × 10^−5^, yielding two standard PRSs with stringent and lenient SNP selection strategies, respectively. To ensure a fair comparison, the elastic net, random forest and XGBoost PRSs were trained on the same two sets of SNPs selected by the C+T procedure using the training data, yielding two PRSs for each method. Hyperparameter tuning followed the same procedure as in the simulation study, and the definition of each PRS was kept identical.

The fitted PRS models were applied to the independent test set to generate PRSs for each individual. Predictive performance was assessed by fitting logistic regression models with ischemic heart disease as the outcome and each PRS, age, sex and the first 10 genetic principal components as predictors. Performance was evaluated using Nagelkerke *R*^2^, AUROC, AUPRC and Brier score. To quantify the incremental predictive value of each PRS, we additionally fitted a baseline model by excluding PRS, and reported the difference between the joint and baseline models for each performance metric. Bootstrapped 95% confidence intervals were calculated for both the joint performance metrics and their incremental differences relative to the baseline model.

## 3. Results

### 3.1 Simulation

Figure 2 presents *R*^2^ of the four PRS methods for the quantitative phenotype across varying proportion of interaction genetic variance *λ*_I_, number of interactions *n*_int_ and sample sizes *n*. The dashed horizontal line represents the “oracle reference”, corresponding to the theoretical upper bound of *R*^2^ under the data-generating mechanism (the broad-sense heritability *H*^2^).

**Figure 2.**
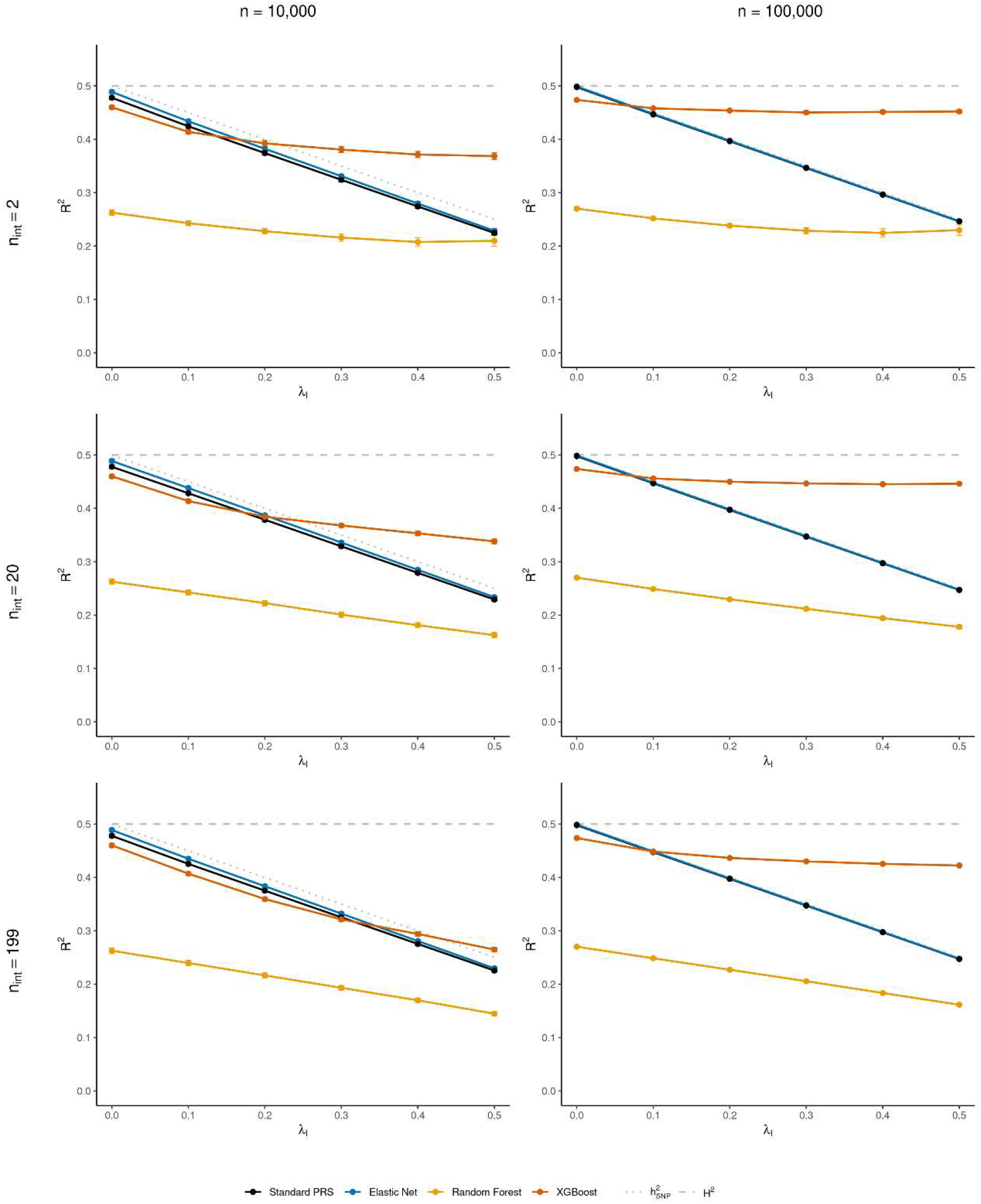
The *R*^2^ of the four PRS methods for the quantitative phenotype across varying proportion of interaction genetic variance *λ*_I_, number of interactions *n*_int_ and total sample sizes *n*. Dotted and dashed grey lines represent additive SNP heritability and broad-sense heritability, respectively.

As *λ*_I_ increased, *R*^2^ of the standard PRS and elastic net decreased approximately linearly, closely tracking the theoretical upper bound of linear models based on effect allele counts, namely the additive SNP heritability. XGBoost also exhibited a decline in *R*^2^ as *λ*_I_ increased, but at a substantially slower rate. Consequently, although XGBoost achieved slightly lower *R*^2^ than the standard PRS and elastic net when *λ*_I_ was low, it outperformed both methods when *λ*_I_ exceeded a configuration-dependent value. Random forest likewise showed slower decrease in *R*^2^ with increasing *λ*_I_ compared to the two linear methods, but consistently achieved the lowest *R*^2^ across all simulation scenarios.

The declining rate of *R*^2^ from XGBoost as *λ*_I_ increased depended strongly on both the concentration of interaction effects *n*_int_ and the sample size *n*. More SNP interactions and larger sample sizes both reduced the declining rate, causing XGBoost to outperform the standard PRS and the elastic net at progressively smaller values of *λ*_I_. For example, at *n* = 10,000, the crossover occurred at approximately *λ*_I_ = 0.16, 0.2, 0.33, when *n*_int_ = 2, 20, 199, respectively. At *n* = 100,000, the correspondingly crossover values reduced to *λ*_I_ = 0.08, 0.085, 0.1. In contrast, *R*^2^of the standard PRS, elastic net and random forest showed little sensitivity to either the concentration of interaction effects or the training sample size. Increasing the sample size from 10,000 to 100,000 yielded only modest improvements in *R*^2^ for all three methods.

Overall, XGBoost substantially outperformed the standard PRSs only when the genetic architecture involved a large proportion of interaction genetic variance and interaction effects from relatively few SNP combinations, and large individual-level datasets were available.

Figure 3 shows the results for the binary phenotype from simulated data. The dashed horizontal line represents the “oracle reference”, corresponding to the highest achievable Nagelkerke *R*^2^ under the simulated liability-threshold model.

**Figure 3.**
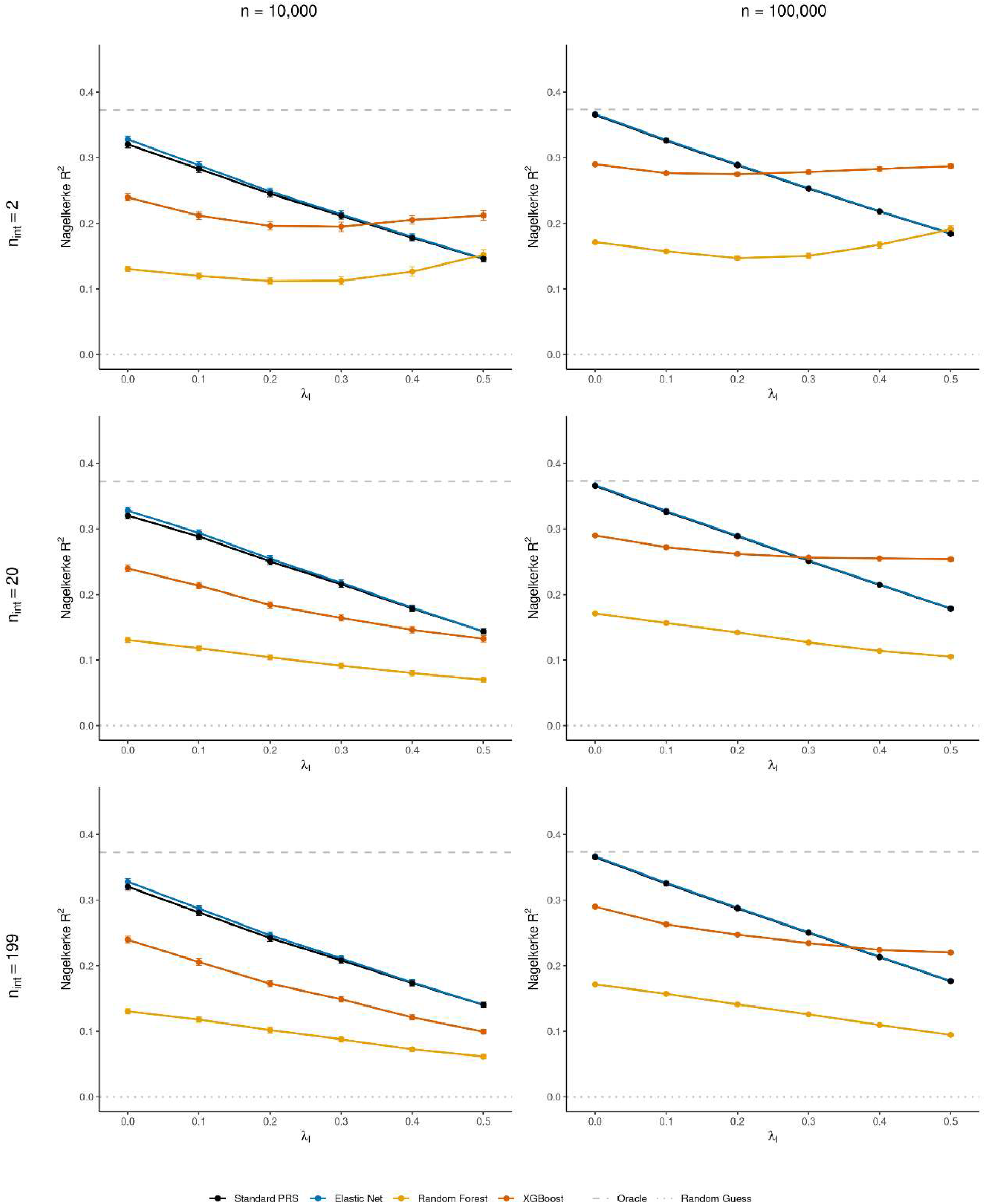
The Nagelkerke *R*^2^ of the four PRS methods for the quantitative phenotype across varying proportion of interaction genetic variance *λ*_I_, number of interactions *n*_int_ and total sample sizes *n*.

Similar patterns were observed to those for the quantitative phenotype. As the proportion of interaction genetic variance increased, the Nagelkerke *R*^2^of the standard PRS and elastic net declined approximately linearly, whereas random forest and XGBoost showed a substantially slower decline. Consequently, XGBoost outperformed the linear methods and stayed closest to the “oracle reference” when interaction genetic variance was sufficiently large, number of interactions was small, and the sample size was large. Random forest consistently achieved the lowest Nagelkerke *R*^2^, with only modest improvement over the standard PRS and elastic net when *λ*_I_ = 0.5 and *n*_int_ = 2. Notably, XGBoost generally required a larger proportion of interaction genetic variance to the linear methods, compared to the quantitative phenotype. Similar conclusions were obtained for AUROC, AUPRC and the Brier score (Supplementary Material Figures S1–S3). Computational costs also increased substantially for the nonlinear methods, particularly random forest and at the larger sample size, with detailed computational times reported in Supplementary Material Table S2.

### 3.2 UK Biobank Data Analysis

The analysis was performed on a case-control cohort of 336,793 UK Biobank participants. The training set comprised 32,847 cases and 236,587 controls, while the test set comprised 8,212 cases and 59,147 controls. We performed a GWAS for ischemic heart disease in the training set, where 9,725,230 SNPs were included in the association analysis. The genomic inflation factor was 1.19, whereas the LD score regression intercept and attenuation ratio were 1.03 and 0.11, respectively, indicating most of the inflation was attributable to polygenicity. Using a genome-wide significance threshold of *P* < 5 × 10^−8^, we identified 45 loci based on the definition from the Global Biobank Meta-analysis Initiative^37^.

Following the C+T procedure, 32 and 162 SNPs were retained under the *P* < 5 × 10^−8^and *P* < 1 × 10^−5^ thresholds, respectively. Both SNP sets were used to construct the standard, elastic net, random forest and XGBoost PRSs.

Figure 4 compares the incremental predictive performance of the four PRS methods relative to the baseline model including age, sex and the first 10 genetic principal components. Across all four performance metrics, the standard PRS, elastic net and XGBoost provided comparable improvements in predictive performance, with largely overlapping 95% confidence intervals. In contrast, random forest consistently yielded smaller gains in Nagelkerke *R*^2^, AUROC and AUPRC, and a smaller reduction in Brier score. Similar patterns were observed under both SNP-selection thresholds (*P* < 5 × 10^−8^ and *P* < 1 × 10^−5^). Overall, these results indicate that, for predicting ischemic heart disease in the UK Biobank, neither elastic net nor XGBoost led to substantial improvements in predictive performance over the standard PRS using C+T, whereas random forest consistently underperformed other methods.

**Figure 4.**
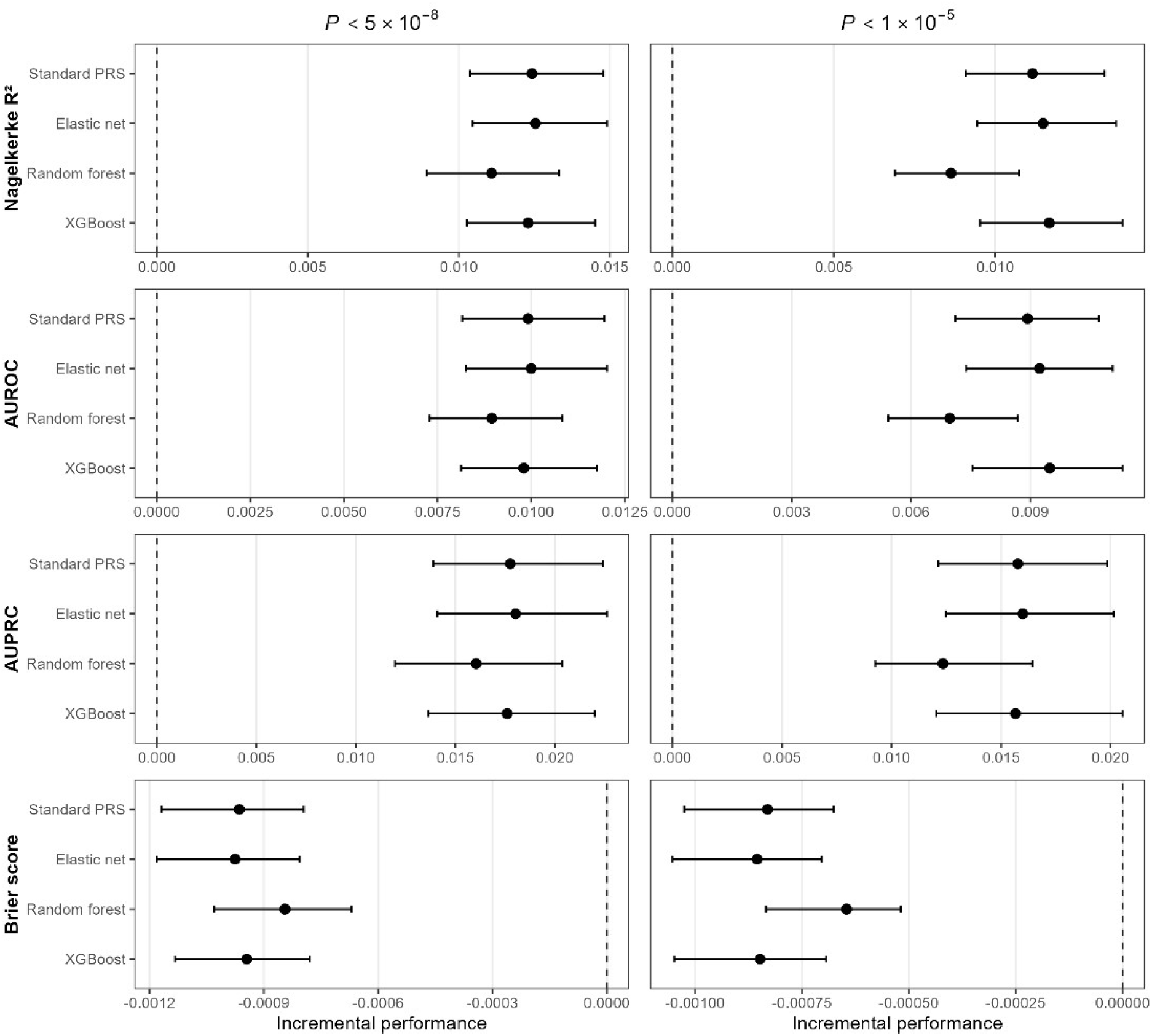
The incremental predictive performance of the four PRS methods relative to the baseline model including age, sex and the first 10 genetic principal components. The negative incremental Brier scores correspond to improved predictive performance.

## 4. Discussion

In this study, we investigated when nonlinear machine learning models, particularly random forest and XGBoost, are expected to improve polygenic risk prediction over standard PRSs, through theoretical derivations, simulation studies and a real-data application.

Theoretically, we showed that standard PRS methods can implicitly capture variance due to interaction genetic effects through their contributions to marginal effects, and therefore may lose less information under interaction genetic architectures than is commonly assumed. This reiterates the distinction between statistical variance components and the underlying genetic architecture illustrated by Huang and Mackay^38^. Specifically, the marginal SNP effects from an additive GWAS are statistical projection coefficients onto the linear space spanned by additive genotype coding (effect allele counts), rather than the underlying additive genetic effects. Consequently, the additive genetic variance is the variance of the best linear predictor based on additive genotype coding and should not be interpreted as the variance arising exclusively from additive genetic effects. The proportion of phenotypic variance explained by standard PRSs is therefore bounded above by the additive SNP heritability, with some LD-aware methods capable of approaching this limit at a rate of *O*(*N*^−1^) as the GWAS sample size *N* increases^10,20^. Further gains in predictive performance of single-trait single-ancestry PRSs may therefore require methods that can exceed this theoretical limit by capturing genetic variation outside the linear space spanned by additive genotype coding. Random forest and XGBoost are such examples that can, in principle, capture any genetic architecture and are therefore only bounded by the higher limit of broad-sense heritability. However, realising this potential is subject to an additional challenge of bias–variance trade-off. Thus, a higher theoretical limit does not necessarily translate into superior predictive performance in finite samples. The practical advantage of nonlinear machine learning models over standard PRSs depends on how much predictive information remains outside the linear space spanned by additive genotype coding and whether it can be learned robustly from finite training samples.

Our simulation study further identified conditions under which XGBoost outperformed the standard PRS. This extends findings from existing studies that XGBoost can improve prediction over standard PRSs for several phenotypes^21,25^. Specifically, three factors influenced whether XGBoost can provide additional predictive benefit. First, the proportion of interaction genetic variance had to be sufficiently large to leave meaningful predictive information beyond that already captured by standard PRSs. Second, interaction effects needed to be concentrated among relatively few SNP combinations. For a fixed interaction genetic variance, this increases the magnitude of individual interaction genetic effects, making them easier to detect and estimate. Third, sufficiently large training sample sizes, were required to learn these interactions reliably, reflecting the substantial data requirements of flexible nonlinear models. Together, these findings reinforce our theoretical argument that the performance of XGBoost depends not only on the amount of interaction genetic variance, but also on whether interaction effects generate sufficiently strong and learnable predictive signals that cannot be captured by standard PRSs.

Random forest consistently underperformed the standard PRS in our simulations, which echoes a previous comparison of these methods for coronary artery disease^26^. A likely explanation is that the greedy recursive splitting used by random forests can make interactions difficult to recover reliably in high-dimensional genetic data, where only a small subset of SNP combinations carries interaction effects. Furthermore, despite random feature and sample selection, individual trees may remain correlated in genetic data, thus limiting the variance reduction achieved through averaging. In contrast, XGBoost builds trees sequentially, with each tree fitted to the residual errors of the previous ensemble, allowing it to progressively refine complex interaction patterns and making it more effective when strong interaction signals are present. Elastic net provided virtually no improvement over the standard PRS across all simulation scenarios, suggesting that using individual-level genotype data alone offers little benefit over standard PRS methods that use GWAS summary statistics when the prediction model remains linear. The improvements were primarily attributable to nonlinear modelling.

Previous studies comparing nonlinear machine learning methods with conventional PRSs have generally reported modest or inconsistent improvements, with some reporting little or no advantage over linear PRSs^23,25–28^ and others observing improvements for selected traits^14,21–25^. Our theoretical and simulation findings provide an explanation for these heterogeneous findings. Rather than expecting nonlinear models to outperform standard PRSs universally, our results suggest that their advantage depends critically on the genetic architecture (proportion of interaction genetic variance and number of interactions) of the phenotype, and the available training sample size.

Our UK Biobank analysis provided a real-world example that neither the elastic net nor XGBoost meaningfully improved predictive performance over the standard PRS for ischemic heart disease, whereas random forest again performed worst, under both the genome-wide significance threshold and the more lenient SNP inclusion threshold. Together with the above theoretical and simulation results, this suggests that the genetic architecture of ischemic heart disease and the available training data may not provide the conditions required for these nonlinear machine learning methods to outperform standard PRSs. However, an advantage of XGBoost may emerge with larger training datasets or for phenotypes with genetic architectures more favourable to nonlinear machine learning methods.

A major strength of this study is the integration of theoretical derivations, controlled simulation experiments and real-data analyses within one framework. Rather than simply comparing predictive performance across methods, we provide a theoretical explanation for why standard PRSs can capture part of the predictive signal generated by genetic interactions and identify the conditions under which nonlinear machine learning methods are expected to provide additional benefit. This framework helps reconcile previously inconsistent findings in the literature and provides a conceptual basis for interpreting future comparisons between standard and nonlinear machine learning PRS methods.

In practice, it remains difficult to determine a priori whether nonlinear machine learning methods are likely to improve prediction over a standard PRS. For a given phenotype, the proportion of interaction genetic variance provides an initial indication of the potential for nonlinear machine learning to improve prediction, and can be estimated using Genomic-relatedness-based Restricted Maximum Likelihood (GREML)^39^ from individual-level data. However, our findings suggest that there is no universal threshold above which nonlinear machine learning methods outperform standard PRSs. The practical advantage also depends on the number of underlying interaction effects and the available training sample size. As existing evidence from evolutionary theories, twin studies and GREML suggests that non-additive genetic variance is relatively limited for most complex human phenotypes^40–43^, substantial gains from nonlinear machine learning methods may only emerge when interaction effects are relatively few and sufficiently large training samples are available. This potentially explains why previous applications to complex human phenotypes have generally reported modest or inconsistent improvements over standard PRSs^21,26,27^. However, these observations may not necessarily extend to human molecular phenotypes, such as circulating biomarkers, gene expression and protein abundance.

A limitation of this study is that we focused on two tree ensemble methods, random forest and XGBoost, as representative nonlinear machine learning approaches. Although these are among the most widely used machine learning methods for PRS construction, our conclusions may not apply to other nonlinear machine learning models. For example, other approaches, such as support vector machine, kernel methods, neural networks or graph-based models, may exhibit different bias–variance trade-offs and relative predictive performance to standard PRS. Nevertheless, our study will serve as a precursor for extension to a broader range of machine learning methods in future research.

A further limitation is that all methods were evaluated using the same set of SNPs. This design intentionally isolated the contribution of the prediction model while avoiding confounding from differences in SNP selection. In practice, however, predictive performance depends jointly on SNP selection and model fitting, and the SNP set that is optimal for a standard PRS may not be optimal for a nonlinear machine learning model. Consequently, our conclusions should be interpreted as comparing modelling strategies conditional on a given SNP set. Future work may include investigations of the joint optimisation of SNP selection and nonlinear modelling.

Finally, our conclusions are specific to PRSs used for prognostic prediction of phenotypes. Other applications of PRSs, including Mendelian randomization, genetic-environment interaction and treatment effect heterogeneity, have different objectives. Thus, the benefit of nonlinear machine learning methods relative to standard PRSs in these settings remain to be investigated.

In conclusion, our theoretical, simulation and empirical results collectively showed that standard PRSs built from additive GWAS summary statistics should not be interpreted as capturing only additive genetic effects, and nonlinear machine learning methods do not universally outperform standard PRSs. Rather, their relative performance depends on the genetic architecture of the phenotype and the available training sample size.

Standard PRSs are likely to remain highly competitive across many complex human traits, whereas nonlinear machine learning methods are expected to provide meaningful improvements under specific genetic architectures and with sufficiently large training datasets. These findings provide both a theoretical explanation for the generally modest gains achieved by nonlinear PRSs in previous studies and a framework for identifying settings in which more complex prediction models are most likely to be beneficial.

## Data Availability

All code and insensitive data produced are available online at https://github.com/JZhu919/NonlinearMLvsStandardPRS

https://www.ukbiobank.ac.uk/

## Acknowledgments

This research has been conducted using the UK Biobank Resource under application number [532314]. JZ was supported by the Medical Research Council [grant number: MR/W007428/1]’. AB and HG acknowledge funding support from the National Institute for Health and Care Research (NIHR) School for Social Care Research, on behalf of the NIHR Three Schools’ Dementia Research Programme [NIHR-SSCR-DS07]. HG acknowledges support from the UKRI AI Programme and the Engineering and Physical Sciences Research Council for CHAI (Causality in Healthcare AI Hub) [grant number EP/Y028856/1]. APM acknowledges support from the National Institute for Health and Care Research (NIHR) Biomedical Research Centre: Manchester (NIHR203308) and the British Heart Foundation (BHF) Research Excellence Award to the University of Manchester (RE/24/130017). The views expressed are those of the authors and not necessarily those of the NIHR or the Department of Health and Social Care. The authors would like to acknowledge the assistance given by Research IT and the use of the Computational Shared Facility at The University of Manchester.

## Author contributions

Conceptualization and Methodology: JZ, APM and HG; Software: JZ and AB; Formal Analysis, Investigation, Visualization and Writing – Original Draft: JZ; Writing – Review & Editing: JZ, AB, APM and HG; Supervision: APM and HG.

## Data and code availability

UK Biobank data: https://www.ukbiobank.ac.uk/. The data and code generated during this study are available at https://github.com/JZhu919/NonlinearMLvsStandardPRS.

