## Supplementary Materials for "Genetic Architecture and Sample Size Impact Relative Performance of Nonlinear Machine Learning and Standard Polygenic Risk Scores"

Table S1: Hyperparameter ranges, optimised metric, number of iterations in Bayesian optimisation and R package for elastic net, random forest and XGBoost PRS.

| Method | Hyperparameters | Optimised metric | Iterations | R package |
|---|---|---|---|---|
| Elastic net | alpha = {0, 0.1, 0.2,..., 1}, lambda = data-dependent default | MSE (quantitative) Deviance (binary) | NA | glmnet |
| Random forest | m_try = [1, p], min.node.size = [1, 0.2n], sample.fraction = [0.1, 0.9], n.trees = 500 | R squared (quantitative) Log-loss (binary) | 30 initial iterations + 50 iterations | tuneRanger |
| XGBoost | eta = [0.01, 0.3], max_depth = [1, 6], subsample = [0.2, 1], colsample_bytree = [0.2, 1], min_child_weight = [1, 50], n_rounds = 500 | R squared (quantitative) Log-loss (binary) | 20 initial iterations + 30 iterations | xgboost mlr3mbo |

Table S2: Mean computation time for 100 simulated datasets, averaged across $\lambda_I$ and $n_{int}$ configurations. Standard PRS and elastic net were computed using single thread. Random forest and XGBoost were computed using 36 CPUs. Computation was performed on Intel "Cascade Lake" Xeon Gold 6230 CPU 2.10GHz + 192GB RAM.

| Phenotype | Sample size | Method | Time | Range |
|---|---|---|---|---|
| Binary | 10,000 | Standard PRS | 8 m | 7–10 m |
| Binary | 10,000 | Elastic net | 1 h 09 m | 48 m–1 h 25 m |
| Binary | 10,000 | Random forest | 2 h 44 m | 2 h 23 m–3 h 18 m |
| Binary | 10,000 | XGBoost | 1 h 54 m | 1 h 04 m–2 h 49 m |
| Binary | 100,000 | Standard PRS | 1 h 08 m | 56 m–1 h 19 m |
| Binary | 100,000 | Elastic net | 15 h 34 m | 13 h 14 m–17 h 21 m |
| Binary | 100,000 | Random forest | 58 h 43 m | 40 h 40 m–84 h 18 m |
| Binary | 100,000 | XGBoost | 5 h 42 m | 4 h 28 m–7 h 54 m |
| Quantitative | 10,000 | Standard PRS | 1 m | 1–2 m |
| Quantitative | 10,000 | Elastic net | 11 m | 10–13 m |
| Quantitative | 10,000 | Random forest | 5 h 28 m | 4 h 27 m–6 h 29 m |
| Quantitative | 10,000 | XGBoost | 2 h 13 m | 1 h 40 m–3 h 12 m |
| Quantitative | 100,000 | Standard PRS | 12 m | 12–13 m |
| Quantitative | 100,000 | Elastic net | 2 h 28 m | 1 h 53 m–3 h 07 m |
| Quantitative | 100,000 | Random forest | 133 h 23 m | 110 h 25 m–156 h 46 m |
| Quantitative | 100,000 | XGBoost | 5 h 48 m | 4 h 21 m–6 h 45 m |

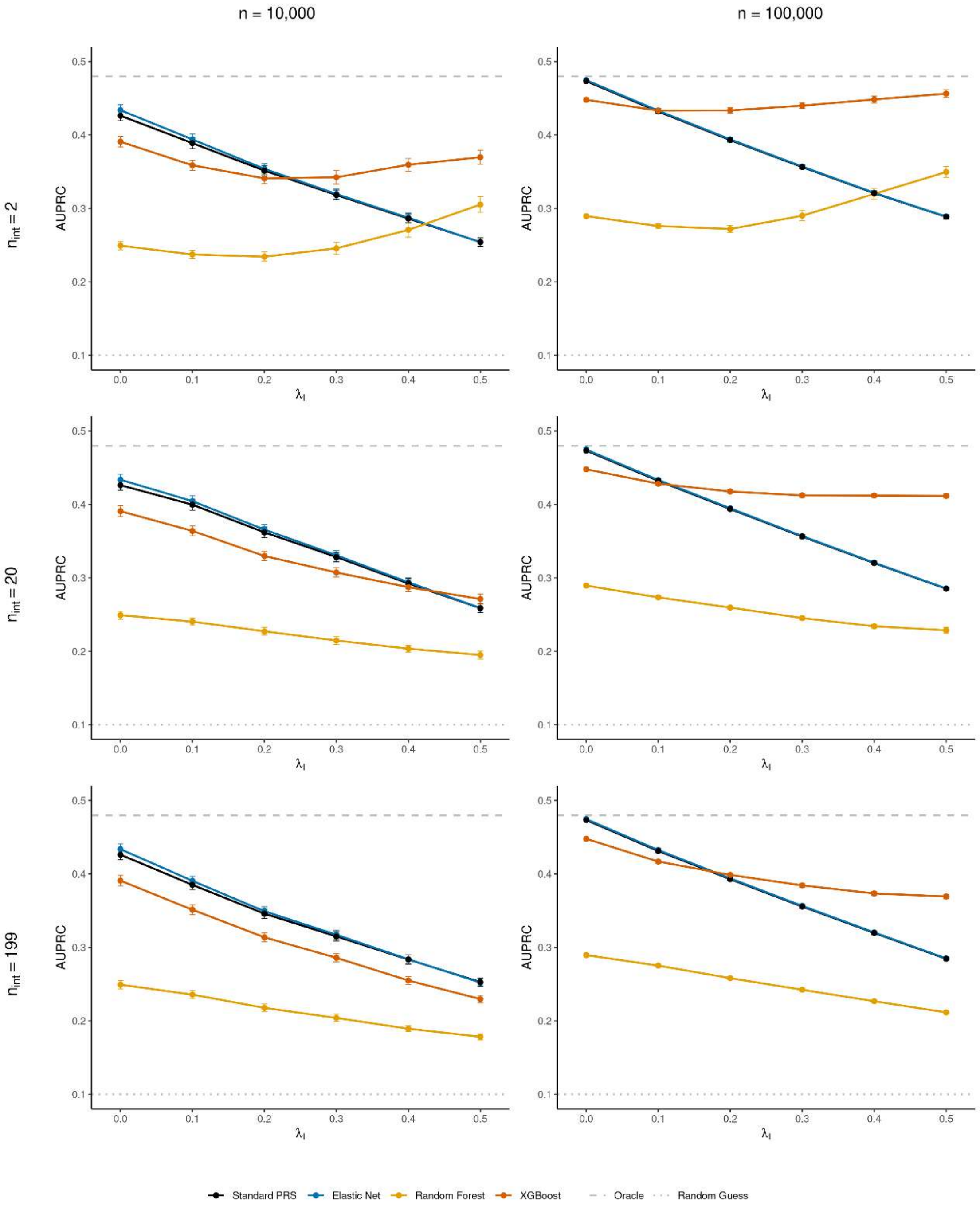


Figure S1. The AUPRC of the four PRS methods for the quantitative phenotype across varying proportion of interaction genetic variance $\lambda_{\mathrm{I}}$, interaction concentrations $n_{\mathrm{int}}$ and sample sizes $n$.

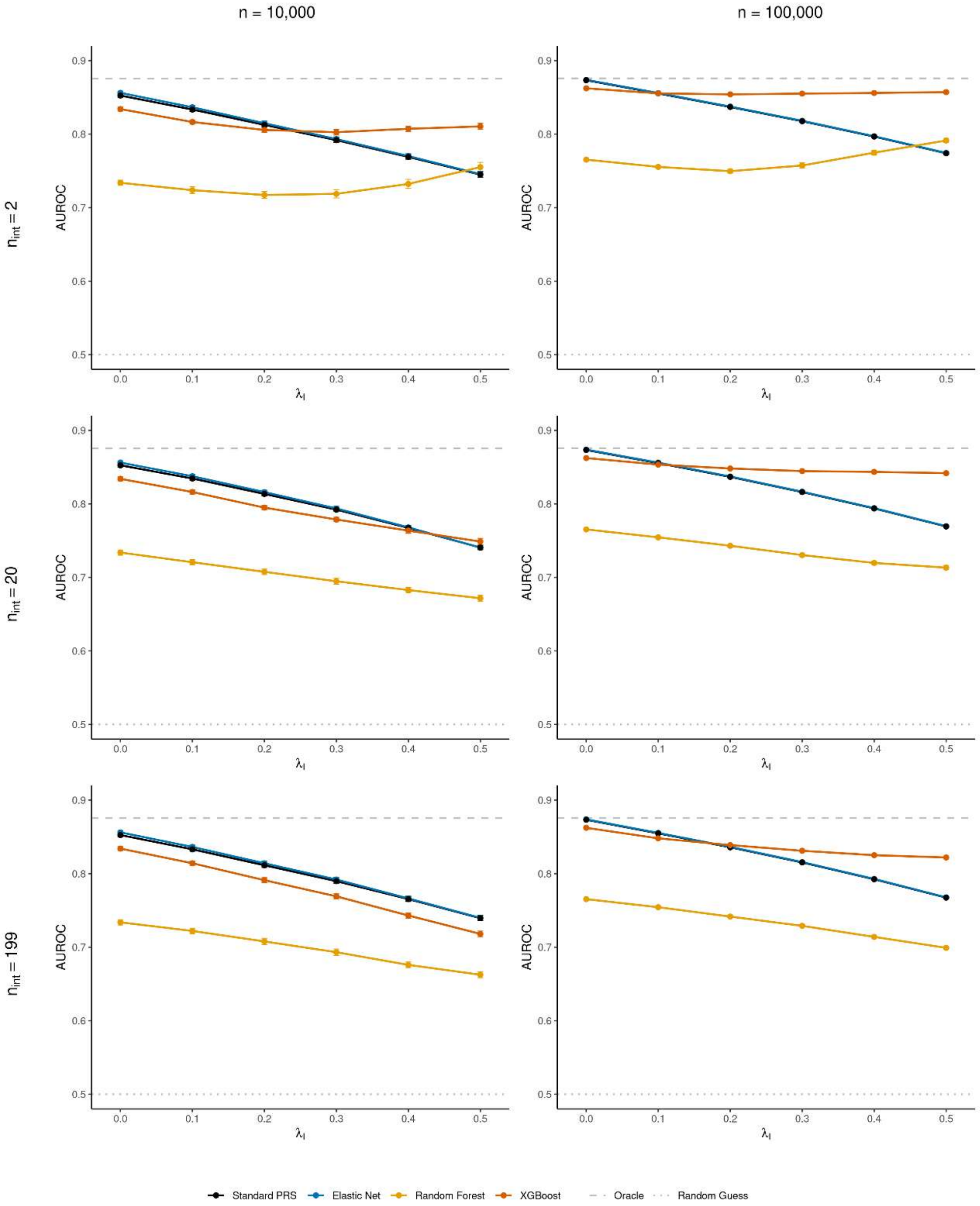


Figure S2. The AURPC of the four PRS methods for the quantitative phenotype across varying proportion of interaction genetic variance $\lambda_{\text{I}}$, interaction concentrations $n_{\text{int}}$ and sample sizes $n$.

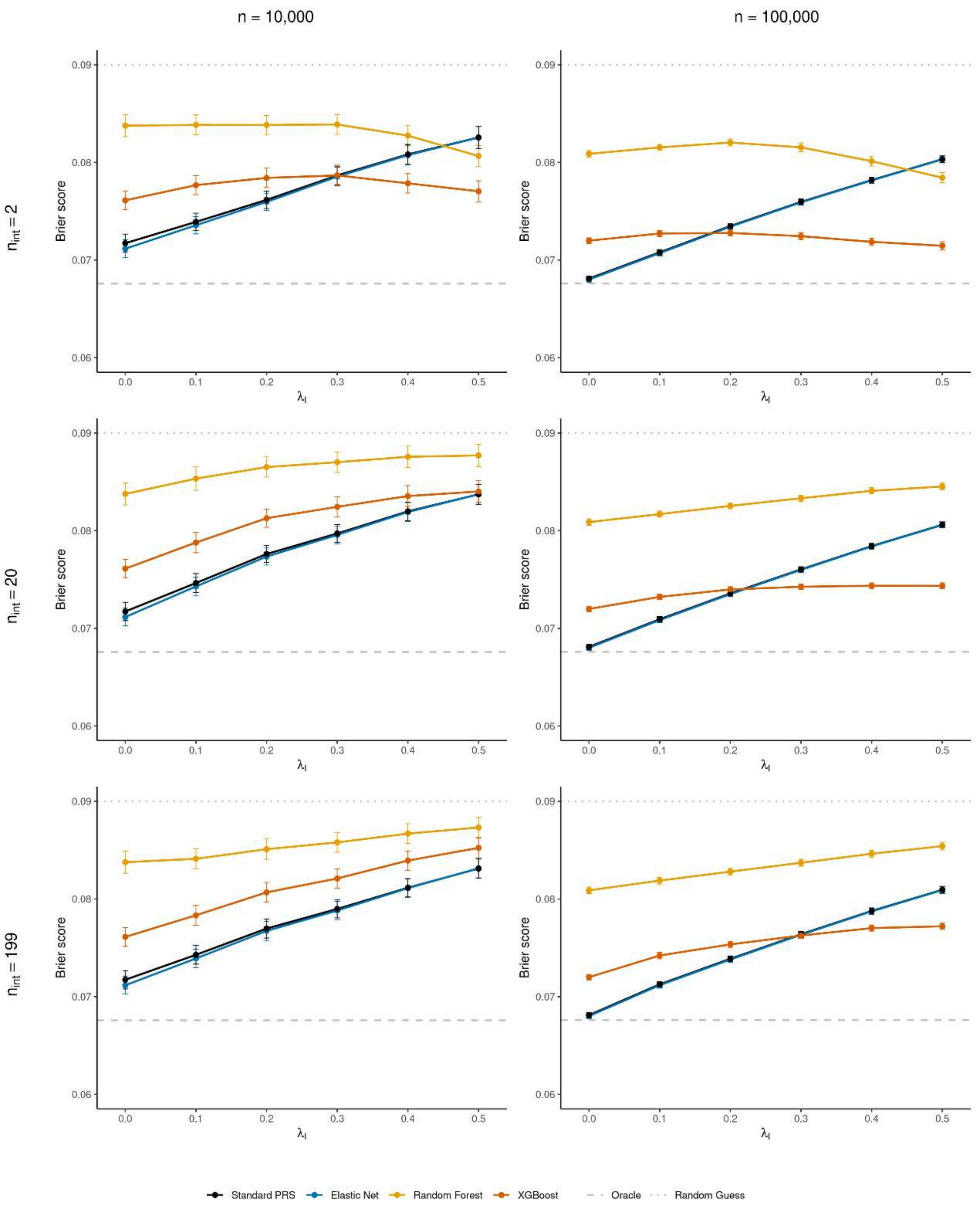


Figure S3. The Brier score of the four PRS methods for the quantitative phenotype across varying proportion of interaction genetic variance $\lambda_{\mathrm{I}}$, interaction concentrations $n_{\mathrm{int}}$ and sample sizes $n$.
